# Automated covid-19 detection from frontal Chest X-Ray images using Deep Learning: an online feasibility study

**DOI:** 10.1101/2020.07.16.20155093

**Authors:** Xavier P. Burgos-Artizzu

## Abstract

**Study design:** to evaluate the performance of Deep Learning methods to detect covid-19 from X-Ray chest images

**Methods:** Chest X-Ray (CXR) images collected from confirmed covid-19 cases in several different centers and institutions and available online were downloaded and combined together with images of healthy patients and patients suffering from bacterial pneumonia found in other online sources. An AI image-based covid-19 classifier was developed and evaluated on the CXR images downloaded.

**Results:** Seven different online data sources were combined for a total of N=16,665 patients (3,156 with covid-19, 2,311 with bacterial pneumonia and 11,198 healthy patients). When half of the patients (N=8,331) where used to train the classifier leaving the other half (N=8,334) for validation, the classifier reached an Area Under the Curve (AUC) for covid-19 detection of 98.6% (detection rate of 91.8% at 1.1% false positive rate). Results were similar for other training/validation splits. AUC was close to 90% even when tested on patients from a source not used to train the classifier.

**Conclusions:** Computer aided automatic covid-19 detection from CXR images showed promising results on a large cohort of patients. The classifier will be made available online for its evaluation. These results merit further evaluation through a prospective clinical study.

## INTRODUCTION

In December 2019, a new coronavirus, was discovered, known as severe acute respiratory syndrome coronavirus 2 (SARS-CoV-2). The disease it causes is called coronavirus disease 2019 (covid-19), a highly infectious respiratory disease^1^. In March 2020, the World Health Organization (WHO) declared the covid-19 outbreak a pandemic.

Symptoms of covid-19 vary in severity from having no symptoms at all to having fever, cough, sore throat, general weakness, fatigue, muscular pain and loss of smell. In the most severe cases it can lead to severe pneumonia and acute respiratory distress syndrome (ARDS) which can lead to death^2^. The time from exposure to onset of symptoms is typically around five days, but may range from two to fourteen days^3^.

The preferred method of diagnosis of covid-19 is by real-time reverse transcription polymerase chain reaction (rRT-PCR) from a nasopharyngeal swab^4^. However, PCR is still prone to errors as covid-19 detector^5^, its precision varying according to the quality of the biological sample obtained^6^. Other tests such as immune response from serological tests are also used but depend on the disease stage and the patient’s immune response to the virus.

Detecting covid-19 from a widely used technique such as Chest X-Ray (CXR) imaging could have its advantages, especially because of the readiness to be applied in most centers (it’s the first-line imaging modality used for covid-19 suspected patients). This is a key factor considering that timely diagnosis of covid-19 is of adamant importance both for the patient’s successful treatment and to control the pandemic spread^7–9^.

However, pathological findings found through CXR cannot be specific to the type of virus causing the pathology. Covid-19 is a viral infection with things in common with other types of viral pneumonia and infections caused by other coronavirus such as influenza, H1N1, SARS or MERS. Nevertheless, in the context of the pandemic, identification and monitoring of lung involvement in potential covid-19 patients can be relevant for clinical treatment and monitoring, especially in intensive care units^10,11^. Moreover, the information provided by such methods could be combined with all the other information regarding the patient (clinical history, symptoms, closeness to other covid-19 cases, etc.) to reach a more informed decision^12^.

In this study, we developed a novel covid-19 classifier from frontal CXR images, using state-of-the-art Artificial Intelligence (AI) methods. We are not the first to attempt this. Since the beginning of the covid-19 outbreak, many researchers have looked at how AI and its many possibilities could help with the pandemic^13^. Covid-19 automated diagnosis from CXR has been proposed before in some prior works^11,14–17^. However, as pointed out by Shi et al ^13^, these studies used a very limited number of covid-19 images, insufficient for a robust evaluation.

In this feasibility study we performed a careful evaluation of the proposed method on a large cohort of patients from different sources online. We evaluate several training/validation scenarios and we directly test if the methods proposed are capable of separating covid-19 from a bacterial infection through the incorporation of bacterial pneumonia patients to the study.

## MATERIALS AND METHODS

### Study design

This was an online retrospective case-control feasibility study. We searched for all frontal (Anterior-Posterior or Posterior-Anterior views) Chest X-Ray images available online. We only downloaded images collected during prior clinical studies and already ordered in the form of proper clinical datasets with anonymized information regarding the patient and verified clinical outcomes. In total, seven different data sources were used. Since all patients were from public databases we were not responsible for collecting patient informed consent and no ethical committee approval was needed.

Three different sources provided CXR images of covid-19 patients: *covid-19 Image Data Collection*^18^, *Covid Data Save Lives*^19^ (*COVIDDSL*) and *PadChest-covid*^20^. After careful revision, we only retained images from patients with a confirmed positive PCR record in the two weeks prior to the image acquisition date.

As stated in the Introduction, it should be possible to distinguish covid-19 infection from bacterial infections. To put these theoretical notions to test, we added CXR images from patients suffering from bacterial pneumonia. We added U.S. National Library of Medicine Tuberculosis Datasets^21^, TB_portals^22^, both containing bacterial tuberculosis CXR images, and QMenta^23^ containing other types of bacterial pneumonia.

From all the above sources we downloaded also all healthy patients, but few were found. Therefore, we added CXR normal images of healthy patients from ChestX-ray8^24^ and PadChest^25^ datasets (5,000 and 6,000 randomly selected patients respectively).

Images from all these sources were already provided after a pre-processing done by the dataset authors: clinical image headers were removed for anonymization, and images converted to either as Portable Network Graphics (PNG) or Joint Photographic Experts Group (JPEG) images. The exception was *COVIDDSL*^19^, which was provided using original Digital Imaging and Communications in Medicine (DICOM) format images. These images were automatically processed in a similar fashion to the other datasets, removing the image header and converting them to PNG format after adequate conversion to 8-bits.

Figure 1 shows some image examples from all sources.

**Figure 1.**
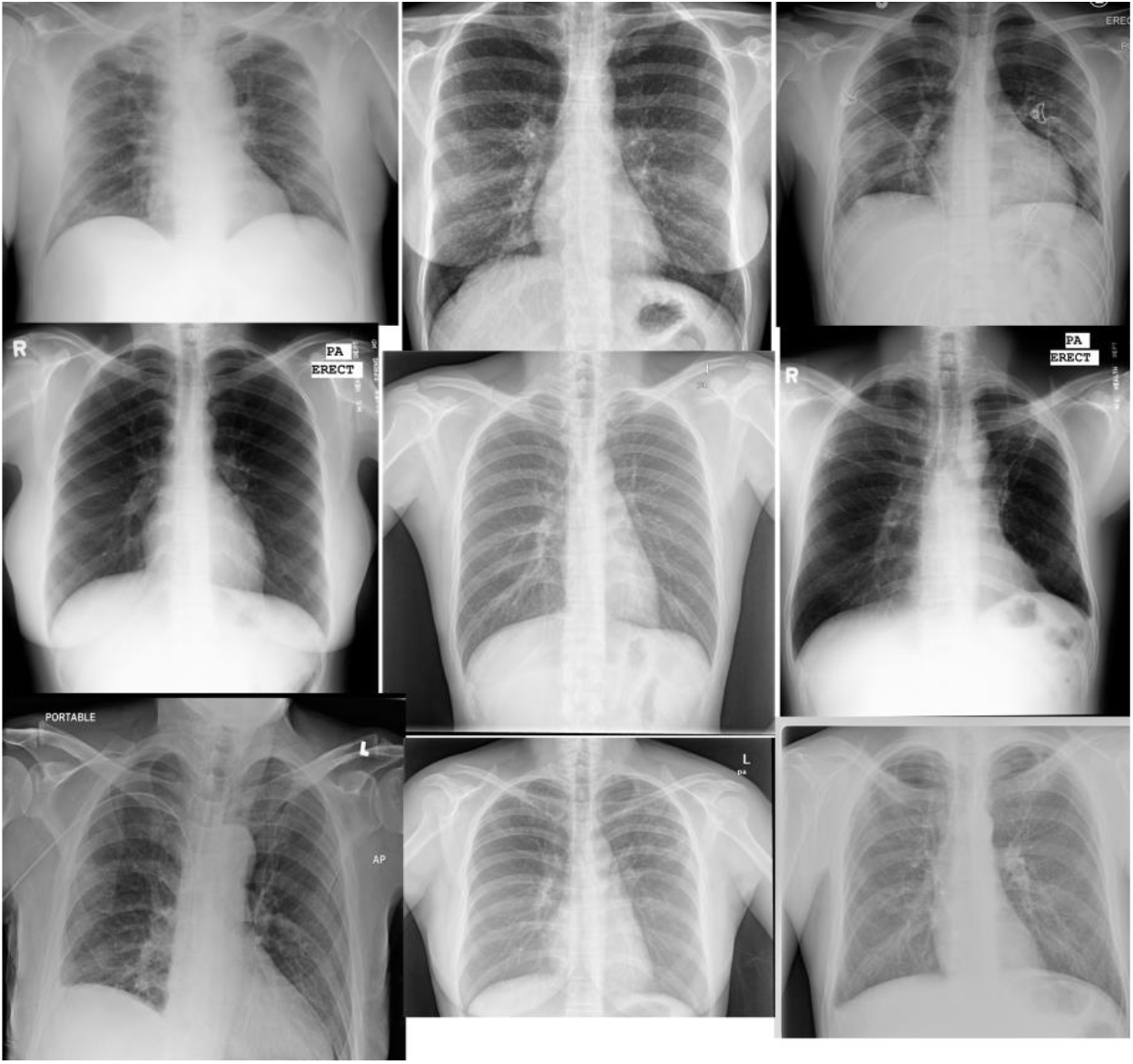
CXR image examples.

### Automated lung segmenter

An automatic lung segmenter was developed using a small fraction of the images downloaded (1,100 images). X.B-A manually delineated both lungs in the CXR image using a Graphical User Interface (GUI) online program. Then, 1,000 images were used to train a Convolutional Neural Network^26^(CNN) and the remaining 100 were left to test the model. More specifically, a variation of a DeepLab CNN model^27^ was used and trained using our AI online platform.

Once trained, the CXR lung segmentation algorithm was applied to on all images, and its output was used to discard all those images where both lungs were not clearly detected and/or if the lungs detected did not take at least 30% of the image pixels. Example automatic lung segmentations are shown in Figure 2.

**Figure 2.**
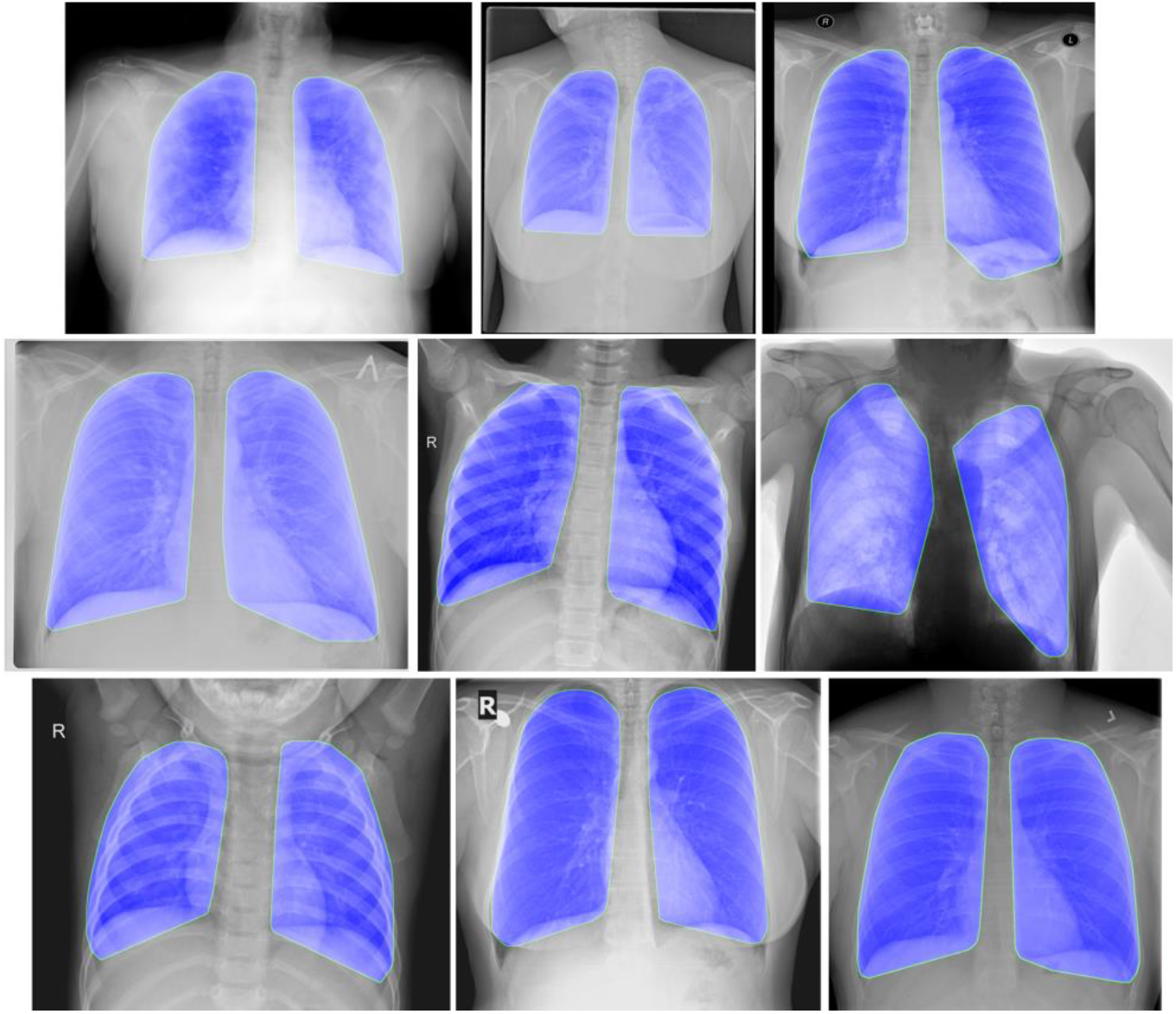
Example automatic CXR Lung segmentation masks.

### Automated CXR image classification

A novel automated image-based CNN classifier was developed. More specifically, we used Inception^28^as backbone architecture. For better performance, we relied on transfer learning. We started by training the original CNN model following the original author’s guidelines on ImageNet^29^. Then, the architecture was changed to be adapted to the study’s task. The first convolution was adapted to work with grayscale images and make sure it only used the image pixels contained in the lung masks (provided by the automated segmenter). Finally, the output layer was changed to output the three class probabilities (healthy, covid-19, bacterial). This adapted net was fully retrained (allowing changes to the entire network) using the study’s training data.

### CNN classifier details

The model was trained using our AI online platform using softmax cross-entropy loss and adam optimizer. 10% of the training set was used as internal validation set. The first training using ImageNet images was performed for 90 epochs with a 5 epoch warm-up. Then, it was trained on our training data for a maximum of 30 epochs, early stopping if loss on validation set was not improved for 5 consecutive epochs. Learning rate was adjusted using a cosine decay starting at 1e-4. Batch size was 64 and weight decay was 0.9. To improve learning, data augmentation was used during training. At each batch, images were randomly mirrored, cropped between 0–20%, translated from 0–10 pixels and rotated between [−15; 15] degrees. No data augmentation was used during testing to assure that output was always the same given the same image.

### Statistical analysis

Characteristics of the study population (patient Gender and Age) were described using percentage and mean and standard deviation respectively. Missing information on these variables was first tested using Little’s test, and then blanks were filled using multiple imputation. Their relationship with clinical outcome was established through t-test hypothesis null testing.

Power sampling^30^ was used to establish the number of patients necessary to validate a classifier with at least 80% detection rate considering the prevalence of the data and error rates of type I and II below 5%. Using that number as bare minimum for validation, many training/validation combinations were evaluated for completeness.

In each experiment, after training, the classifier was deployed and applied to all validation images, outputting full probability scores for each class. If the patient had more than one image taken the same day, we averaged the output probability scores of the classifier and used these numbers as final class probabilities for that patient.

All statistical analysis was then performed using Matlab (Mathworks, USA). The class probability scores were used to compute the class confusion matrix and report global accuracy and average class-accuracy. Then, individual class scores for covid and/or bacterial pneumonia were used to draw Receiver Operator Characteristic (ROC) curves and compute full Area Under the Curve (AUC). Finally, optimal cutoff points (maximizing accuracy) were computed from the ROC and used to report Detection rate, false positive rate (FPR), positive and negative predictive values (PPV and NPV) and positive and negative likelihood ratios (LR+ and LR-). All metrics were calculated together with their 95% Confidence Intervals (estimated using 10 bootstrapping rounds on the validation set).

Further analysis was performed, such as a visual inspection of the CNN saliency maps^31^ for interpretability of results. Then, mistakes of the classifier (incorrect class predictions) were analyzed for possible statistical correlations with patient’s age, gender or origin, using t-test hypothesis null testing.

## RESULTS

### Final data

A total of 30,974 distinct CXR images were found and downloaded. The lung segmenter was applied to all the images, and its output was used to filter images, retaining only those where both lungs were detected and took at least 30% of the image pixels. In total, the segmenter filtered 269(0.8%) bad quality images, which were therefore removed from the study.

Table 1 shows the resulting dataset, with the number of patients their demographics (gender and age) and total CXR images from each source. A total of 16,665 patients were included in the study, with a covid-19 prevalence of 18.9% (3,156/16,665) and a Tuberculosis prevalence of 13.8% (2,311/16,665). In average each patient had 1.8 CXR images available, for a total of 30,705images.

**Table 1.** CXR data used for the study. Data given as: mean (+-std) or n (%).

| Source | Patients | Covid-19 patients | Bacterial pneumonia patients | Age | Male | Images | Images/patient |
| --- | --- | --- | --- | --- | --- | --- | --- |
| *Covid-19 Image Data Collection*<sup>18</sup> | 182 | 173(95.1%) | 0(0.0%) | 58.5(+/-15.4) | 95(52.2%) | 270(0.9%) | 1.5 |
| *COVIDDSL*<sup>19</sup> | 1,719 | 1719(100.0%) | 0(0.0%) | 67.8(+/-15.3) | 1063(61.8%) | 5,158(16.8%) | 3.0 |
| *NIH ChestX-ray8*<sup>24</sup> | 4,983 | 0(0.0%) | 0(0.0%) | 45.2(+/-16.6) | 2672(53.6%) | 11,760(38.3%) | 2.4 |
| *PadChest*<sup>20,25</sup> | 5,938 | 1264(21.3%) | 69(1.2%) | 57.9(+/-17.0) | 2552(43.0%) | 7,165(23.3%) | 1.2 |
| *TB_portals*<sup>22</sup> | 497 | 0(0.0%) | 417(83.9%) | - | - | 1,200(3.9%) | 2.4 |
| *U.S.N.Library of Medicine Tuberculosis Datasets*<sup>21</sup> | 798 | 0(0.0%) | 393(49.2%) | - | - | 798(2.6%) | 1.0 |
| *Qmenta*<sup>23</sup> | 2,548 | 0(0.0%) | 1432(56.2%) | - | - | 4,354(14.2%) | 1.7 |
| **TOTAL** | 16,665 | 3,156(18.9%) | 2,311(13.9%) | 54.3(+/-16.1) | 8,539(51.2%) | 30,705(100.0%) | 1.8 |

**Table 2.** Statistical relationship between Demographics and Covid-19 infection. Data given as: mean (+-std) or n/N (%).

| Variable | Covid-19 patients | Other (healthy or bacterial pneumomina) | t-test p-value |
| --- | --- | --- | --- |
| Male (%) | 1834/3156 (58.1%) | 6705/13509 (49.6%) | 0.00 |
| Age (years) | 65.0 (+-16.1) | 51.8 (+-15.1) | 0.00 |

As reported in prior works, both the gender and age of patients were found to be statistically correlated with respect to covid-19 infection, with older males showing a higher risk of suffering covid-19. This demographical information was not used as input to the classifier.

The reasons are three-fold: 1) this data was not always available for all images, 2) To avoid overfitting due to dangerous data correlations and 3) to test the power of image analysis on its own.

### Main results

The result of the power sampling to establish number of patients necessary to validate the classifier was N=1,314 patients. This represented only 8% of the data. However, for fairness we used a 50%/50% training/validation split respecting original disease prevalence as main experiment. 8,334 patients were used for validation and the remaining (8,331) for training, All 1,100 images used during the development of the lung segmenter were placed in training for fairness.

Figure 3 shows the Confusion Matrix and ROC curves for covid-19 and tuberculosis detection from CXR images using the testing patients (N=8,334). Global accuracy on the three classes (healthy, covid, bacterial pneumonia) was 93.7%, with a 88.9% average class-accuracy (standard deviation=8.1%). Individual AUC was 98.6% (CI 95 +-0.0%) for covid-19 and 98.1% (CI 95+-0.0) for tuberculosis.

**Figure 3.**
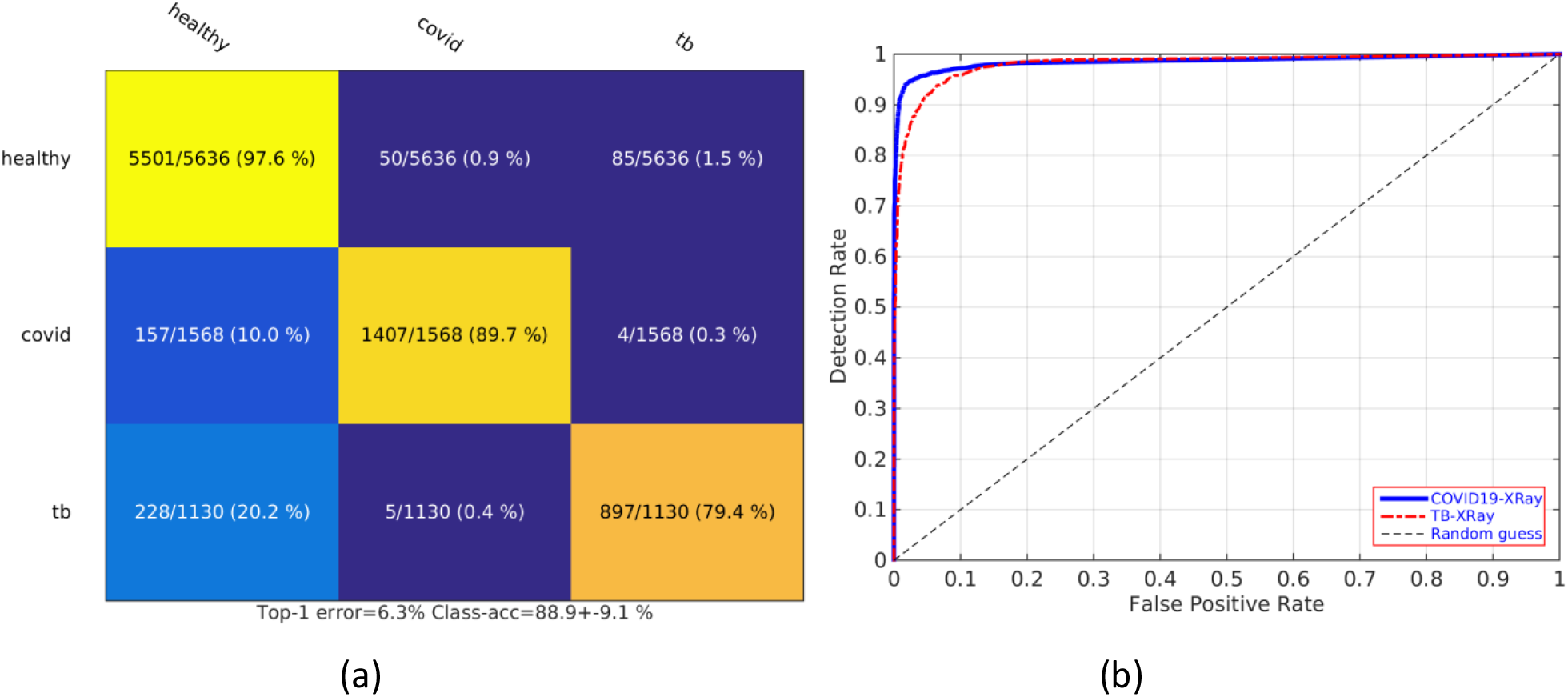
Covid-19 detection results from CXR (N=8,334 Patients, covid-19 prevalence 18.8%, Tuberculosis prevalence 13.5%). (*a*) *Full confusion matrix*. (*b*) *covid-19* and Tuberculosis ROC curves (blue=covid, red=tuberculosis).

Table 3 shows the detailed metric scores at the optimal accuracy cut-off points computed from the ROC curve separately for covid-19 and bacterial pneumonia. In the case of covid-19 the classifier would have been capable of detecting 91.8% (CI 95 +-0.5%) of the infected patients, while incurring in a 1.1% (CI 95 +-0.1%) false positive rate. PPV was 95.2%(CI 95 +-0.5%) and NPV 98.1%(CI 95 +-0.1%). This scenario would have implied that of the 1,568 patients tested that actually had covid-19, the classifier would have successfully detected 1,440, leaving 128 undetected, and on the other hand it would have falsely flagged 72 patients as having covid-19.

**Table 3.** CXR covid-19 & Tuberculosis detection results (N=8,334 Patients, covid-19 Prevalence 18.8%, Tuberculosis Prevalence 13.5%). AUC= Area under the Curve. PPV= Positive Predictive Value. NPV=Negative Predictive Value. LR+= Positive likelihood ratio, LR-=Negative likelihood ratio.

| Disease | Accuracy | F1-Score | AUC | Detection Rate | False Positive Rate | PPV | NPV | LR+ | LR- |
| --- | --- | --- | --- | --- | --- | --- | --- | --- | --- |
| Covid-19 | 8134/8334<br>(97.6 % +-<br>0.0 %) | 93.5<br>% +-<br>0.1 % | 98.6<br>% +-<br>0.0 % | 1440/1568<br>(91.8 % +-<br>0.5 %) | 72/6766<br>(1.1 % +-<br>0.1 %) | 1440/1512<br>(95.2 % +-<br>0.5 %) | 6694/6822<br>(98.1 % +-<br>0.1 %) | 86.3<br>(+-<br>8.2) | 0.1<br>(+-<br>0.0) |
| Tuberculosis | 8015/8334<br>(96.2 % +-<br>0.0 %) | 85.6<br>% +-<br>0.1 % | 98.1<br>% +-<br>0.0 % | 947/1130<br>(83.8 % +-<br>0.6 %) | 136/7204<br>(1.9 % +-<br>0.1 %) | 947/1083<br>(87.4 % +-<br>0.5 %) | 7068/7251<br>(97.5 % +-<br>0.1 %) | 44.4<br>(+-<br>1.7) | 0.2<br>(+-<br>0.0) |

In comparison, the detection of bacterial pneumonia had slightly worse results, with a detection rate of 83.8% (CI 95 +-0.6%) for a false positive rate of 1.9%(CI 95 +-0.1%). PPV was 87.4%(CI 95 +-0.5%) and NPV 97.5%(CI 95 +-0.1%).

Further analysis of results is shown in Supplementary Materials. Errors seemed to be correlated with the patient’s age and gender (older females patients being harder to classify). Finally, saliency maps seemed to indicate that the classifier was focusing on both lungs as a whole.

### Additional experiments

For completeness, we performed two additional experiments using the same initial data presented in Table 1, but varying the amount and type of patients used for training and validation of the CXR classifier. Results are shown in Figure 4.

**Figure 4.**
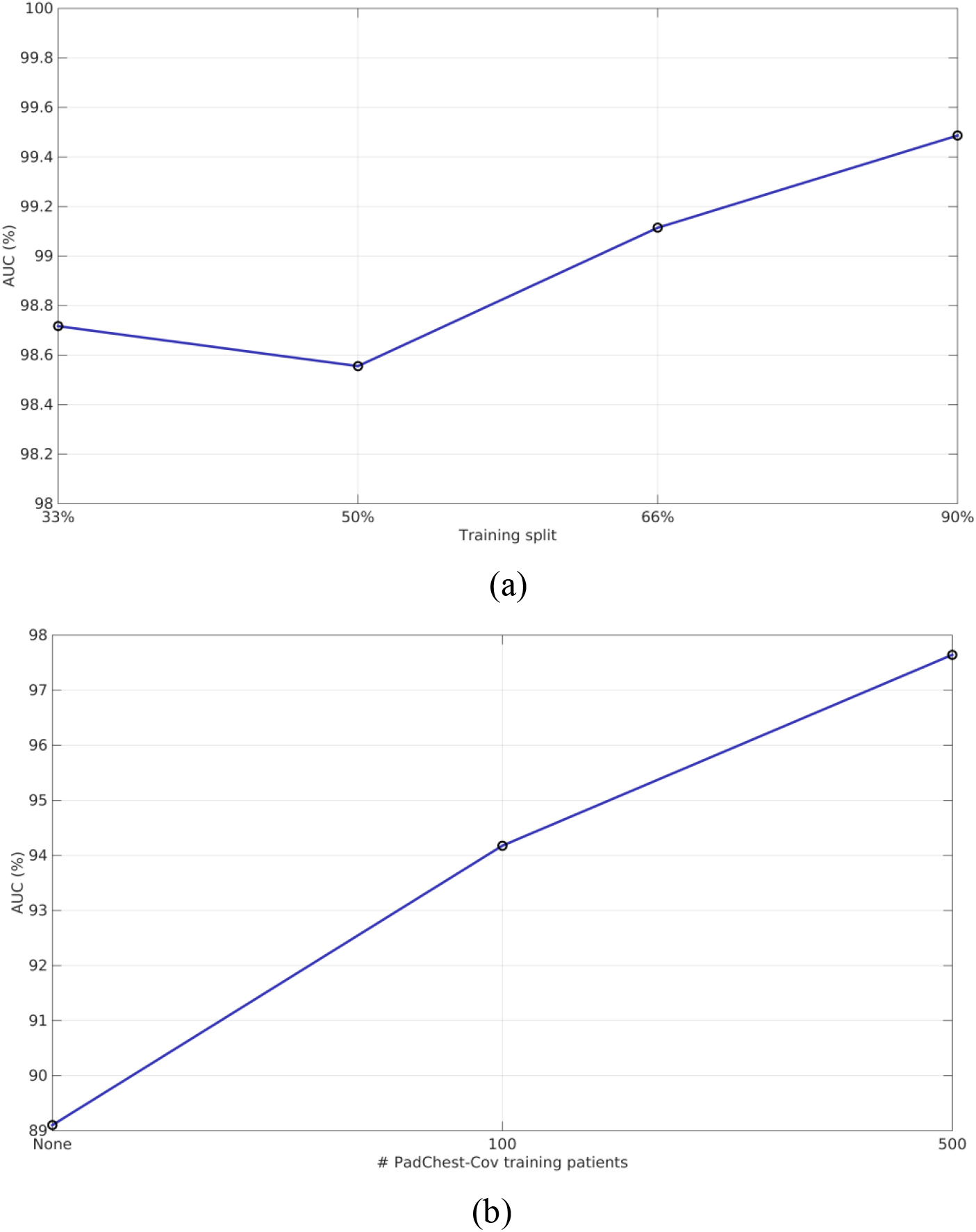
Additional experiments. (a) AUC vs Amount of data used for training [33%, 50%, 66%, 90%]. (b) AUC on PadChest-Covid patients vs amount of PadChest-Covid patients seen during training [None, 100, 500].

First, apart from the 50%/50% training/validation split originally used, we also evaluated how results were affected if other proportions were used. Figure 4(a) shows the results in terms of individual AUC for covid-19 detection on splits using 33%, 50%, 66% and 90% of the patients for training. In general, using more patients for training leads to better performance, as one would expect, but AUC for all cases remains above 98% and differences remain below 1%.

Finally, we designed an experiment to evaluate the covid-19 classifier’s transfer-ability. We analyzed the performance of a classifier trained using as source for covid-19 patients only *CDDSL* and *Covid-19 Image Data Collection* when evaluated on *PadChest-Covid* patients. We simulated both a “hard” transfer (no PadChest-Covid patients were ever used during training) as well as softer scenarios (100 or 500 *PadChest-Covid* patients used for training). Results are shown in Figure 4(b). Blind transfer resulted in an AUC of 89.1%. Adding 100 patients improved AUC to 94.1%, while adding 500 patients improved AUC to 97.6%.

## DISCUSSION

### Main findings

A novel DL classifier to detect covid-19 in CXR images was developed and evaluated on images downloaded online of patients collected from multiple centers and institutions. Results suggest that it is capable of detecting covid-19 afflicted patients through a frontal CXR with high detection rates and low false positive rates. The classifier developed is fully automated since it includes an automatic lung segmenter.

The classifier reached an AUC for covid-19 detection of >98% on the 8,834 patients tested (1,568 of which had covid-19). This translated in a detection rate of 91.8% at 1.1% false positive rate and NPV of 98.1%. It was also able to distinguish the viral infection caused by covid-19 from bacterial pneumonia, for which it obtained an AUC of 98.1%, similar to previously results from large studies on tuberculosis vs other viral infections ^32^.

The classifier showed promising results even when blindly transferred to patients from a center never used for training. Even if this resulted in a 9.5% drop in AUC (from 98.6% to 89.1%), an AUC close to 90% still would not make the classifier useless. And as soon as 100 patients from the “new center” were incorporated into training, AUC went back to numbers close to 95%.

### Clinical implications

Repeated RT-PCR from nasal swabs, the current gold standard for covid-19 detection, is not perfect (no test ever really is). Prior systematic reviews reported varying detection rates between 63% and 98%^6,33^. The main reasons for these variations are the quality of sampling, the stage of the disease the day of the test, the degree of viral multiplication and the different prevalence of the disease in each center^34,35^. Due to these variations, it has been proposed to combine repeated PCR with serology tests, clinical history, history of contact with other covid-19 patients and findings from either chest CT scans and/or X-Ray^12^.

In this study, we evaluated automated methods of covid-19 detection from CXR images on a large pool of patients from different centers. The new AI method developed extracts information from CXR automatically, making the process both easier and quicker compared to the evaluation of images by an expert radiologist.

The high detection rate and NPV reported on the 8,834 patients tested implies that a method such as the one proposed could maybe help in patient management during the covid-19 pandemic, since it could help reduce the number of false negatives. However, the method was tested only on patients in need of clinical care. The degree of affection or symptoms needed before the method is able to detect covid-19 in a patient remains unclear. Nevertheless, considering the many reported limitations of PCR discussed above, reducing false negatives in symptomatic patients through a fast and non-invasive procedure could be useful.

### Strengths

This study has several strengths. We have performed the study combining patients from several sources, each of which was in turn constructed using patients from different centers. This means that the data used is inherently multi-center and multiple equipment and operators were responsible for the acquisition of the images, giving more credibility to the results. Furthermore, we have evaluated the impact on performance of different patient’s training-validation splits and the relationship between patient’s variables and classifier’s mistakes. Moreover, we evaluated the classifier in a real transfer scenario, showing how performance dropped and by how much. Last but not least, for the first time as far as we know, we demonstrated that AI methods seem to be able to distinguish between covid-19 and bacterial infections (covid-19 vs bacterial pneumonias).

### Limitations

We acknowledge a number of limitations. We were not involved in data acquisition; we trusted the data as it came and weren’t able to double-check clinical outcomes of the images.

However, these were public sources collected in large clinical institutions and widely used. Another limitation is the varying prevalence regarding covid-19 patients found in the datasets used, and the need to incorporate random controls from other sources. Finally, all covid-19 patients evaluated were patients in need of clinical care; it is unclear whether the methods evaluated could be used to detect asymptomatic patients. A prospective clinical study is needed to evaluate the limits of the methods proposed.

### Conclusions

An AI covid-19 classifier from CXR images showed promising results. These methods could help to manage patients better in case access to PCR testing is not possible or to detect patients missed in a first round of PCR testing. It will be made available online for its evaluation. These results merit further evaluation and we hope to start large studies with collaborating centers.

## Data Availability

All data used comes from external open research sources available online. The rest of data is available in the main text. The final X-Ray model will be provided online at https://www.quantuscovid19.org

## Author contributions

X.B.-A. was the sole author of this work

## Details of Ethics Approval

(Stated in Methods’ section) Since all patients were from public databases we were not responsible for collecting patient informed consent and no ethical committee approval was needed.

## Data availability

All images and clinical data associated were from public databases, and the final model will be made available online at www.quantuscovid19.org

## Supp Material for paper

Supp Figure 1 shows example correct positive covid-19 detections, with the saliency maps^1^ of the CNN next to the original image. The CNN appears to be focusing on alterations in both lungs.

**Supplementary Figure 1.**
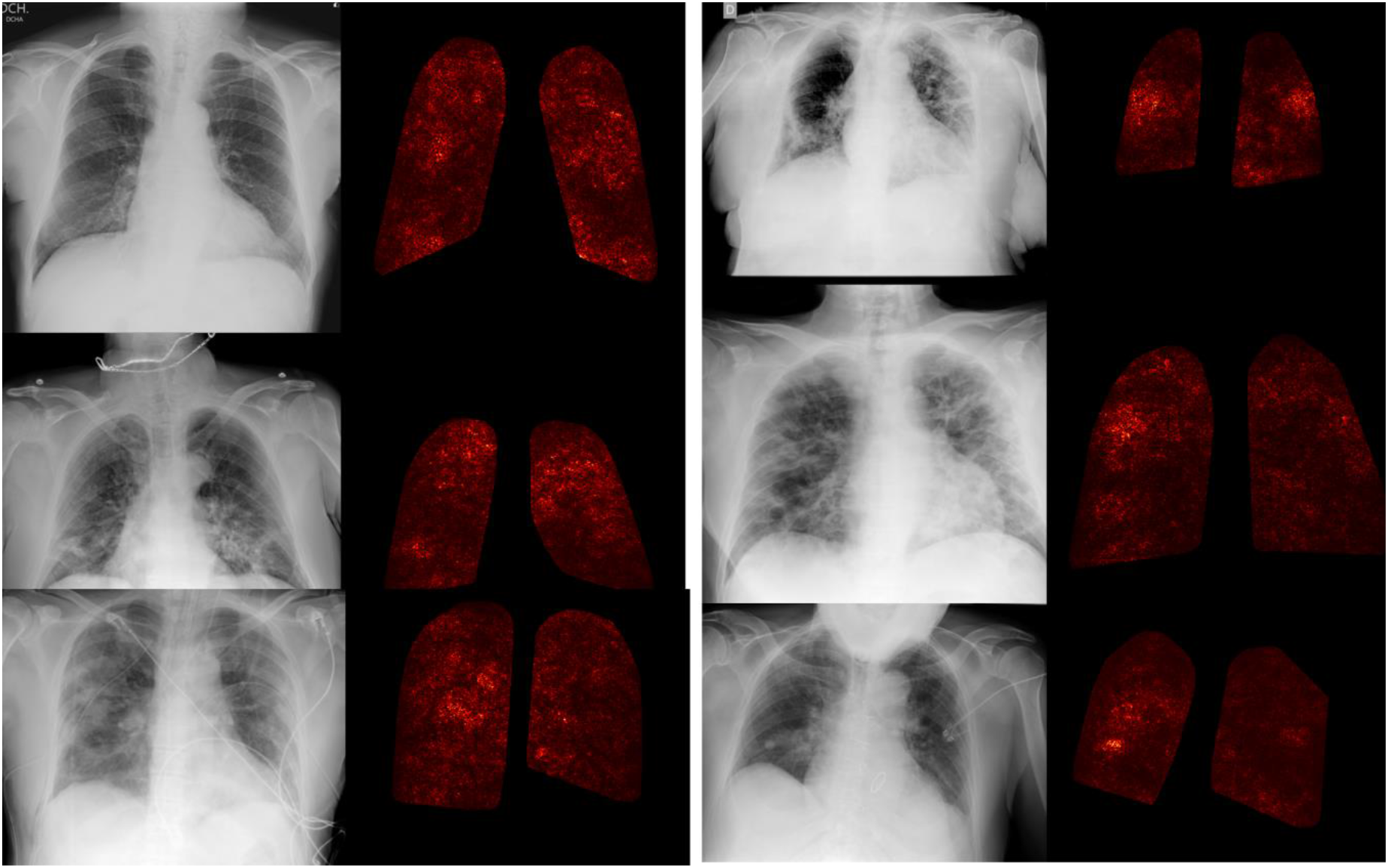
Saliency maps^1^ of classifier on correctly classified covid-19 images.

Supp Table 1 shows the statistical relationship between available patient variables (Gender, Age, Origin) and the covid-19 classification. Male and origin do not seem to affect classifier’s performance. However, age seems to be slightly correlated, with older patients being more difficult to predict.

**Supp Table 1.** Statistical relationship between covid-19 classification and patient’s variables. Data shown as Mean (+-std). p-value computed using a 2 sample t-test.

| Variable | Correct classification | Incorrect classification | t-test p-value |
| --- | --- | --- | --- |
| Male | 4197/8134(51.6%) | 88/200(44.0%) | 0.03 |
| Age (years) | 54.0(+16.2) | 57.2(+17.5) | 0.01 |

